# Structural adjustment programmes and infectious disease mortality

**DOI:** 10.1101/2021.03.12.21253462

**Authors:** Elias Nosrati, Jennifer B. Dowd, Michael Marmot, Lawrence P. King

## Abstract

International financial organisations like the International Monetary Fund (IMF) play a central role in shaping the developmental trajectories of fiscally distressed countries through their conditional lending schemes, known as ‘structural adjustment programmes’. These programmes entail wide-ranging domestic policy reforms that influence local health and welfare systems. Using novel panel data from 187 countries between 1990 and 2017 and an instrumental variable technique, we find that IMF programmes lead to over 70 excess deaths from respiratory diseases and tuberculosis per 100,000 population and that IMF-mandated privatisation reforms lead to over 90 excess deaths per 100,000 population. Thus structural adjustment programmes, as currently designed and implemented, are harmful to population health and increase global infectious disease burdens.

## Introduction

In the wake of the COVID-19 pandemic, much scientific effort has been devoted to better understanding the institutional determinants of communicable disease control. We seek to contribute to this understanding by assessing the role of international multilateral organisations in shaping communicable disease burdens across the world. As one of the world’s leading international financial institutions, the International Monetary Fund (IMF) is uniquely positioned to shape the developmental trajectories of fiscally distressed nations through its conditional lending schemes, known as ‘structural adjustment programmes’. In particular, the Fund plays a pivotal role in shaping state capacities to manage pandemics by moulding the institutional infrastructure of local health and welfare systems (1).

Previous research has shown that the IMF-sponsored pursuit of short-term economic goals at the expense of long-term public investments by financially constrained governments can undermine local health systems via fiscal austerity and rapid privatisation reforms. For instance, the curtailing of healthcare coverage and sapping of resources from primary care can weaken the quality of care provision (2, 3). More generally, IMF-mandated policy reforms — known as ‘conditionalities’ — have been associated with reduced state capacities and declining population health (4–6). However, with the exception of a few case studies (7, 8), we are not aware of any prior systematic investigations of the causal relation between the IMF’s structural adjustment programmes and communicable disease burdens.

A separate literature has shown that the rapid privatisation of state-owned enterprises can damage population health by increasing unemployment and social insecurity, as well as eroding the public provision of various social goods, including access to healthcare (9, 10). However, research on the nexus between privatisation and mortality through the lens of structural adjustment remains scarce, and virtually nothing is known about how IMF-mandated privatisation reforms affect disability and death burdens from communicable diseases. Our paper fills this gap by using previously unavailable data and a compound instrumental variable to derive statistically robust effect estimates.

## Methods

Our outcome variable is the country-level age-standardised mortality rate from respiratory infections and tuberculosis (TB) per 100,000 population between 1990 and 2017. Taken together, these causes of death make up category A.2 within the framework of the Global Burden of Diseases, Injuries, and Risk Factors Study, the methodology of which is employed to generate comparable age-standardised mortality metrics across country-years (11). We employ two sets of treatment variables to assess the effects of structural adjustment (12). On the one hand, we use a dichotomous indicator of whether a country is under an IMF programme to estimate an overall average treatment effect of IMF intervention. On the other hand, to further probe the specific nature of structural loan conditions and their relation to the outcome variables, we assess the role of IMF-mandated privatisations of state-owned enterprises.

Our economic control variables are gross domestic product (GDP) per capita, measured in constant 2010 US dollars (13), a binary financial crisis indicator (14), and foreign reserves in months of imports (13). Our political control variables include a general democracy index (15) and a more refined measure of egalitarian democracy (16), a coup d’état indicator (17) as a measure of political instability, and United Nations General Assembly (UNGA) voting alignment with the G7 countries (18). The latter variable is construed as a proxy for geo-strategic alignment and is known to be predictive of IMF programme participation and potentially of the types of conditionalities received by borrowing countries (19). We also control for average years of completed education in the female population aged 25–29 (20). Finally, to test the hypothesis according to which much of the proposed impact propagates through health systems, we control for the number of hospital beds per 1,000 population (13). Descriptive statistics are shown in Table 1.

**Table 1:** Descriptive statistics

| STATISTIC | N | MEAN | ST. DEV. | MIN | MAX |
| --- | --- | --- | --- | --- | --- |
| Communicable disease mortality rate | 5,460 | 96 | 103 | 5 | 613 |
| IMF programme | 5,165 | 0.3 | 0.5 | 0.0 | 1.0 |
| IMF-mandated privatisation | 5,131 | 0.1 | 0.5 | 0.0 | 12 |
| GDP per capita | 4,733 | 11,561 | 17,103 | 164 | 111,968 |
| Financial crisis | 5,236 | 0.1 | 0.2 | 0.0 | 1.0 |
| Foreign reserves | 3,932 | 4.4 | 4.7 | 0.002 | 79 |
| Democracy | 4,909 | 3.5 | 6.6 | -10 | 10 |
| Egalitarian democracy | 4,097 | 0.4 | 0.2 | 0.03 | 0.9 |
| Coup d'état | 4,396 | 0.02 | 0.1 | 0.0 | 1.0 |
| UNGA voting alignment | 5,012 | -1.6 | 1.0 | -3.9 | 1.4 |
| Mean years of female education | 5,404 | 8.9 | 4.0 | 0.5 | 16 |
| Hospital beds per 1,000 population | 2,694 | 4.1 | 3.1 | 0.01 | 20 |

NOTES: The mortality rate from communicable diseases is age-standardised per 100,000 population and refers to deaths from respiratory infections and tuberculosis. The second column lists the number of observed country-years. The privatisation variable counts the total number of privatisation conditionalities imposed on a borrowing country by the IMF. The general democracy index ranges from -10 to 10. The egalitarian democracy index ranges from 0 to 1.

We posit the following data-generating process:

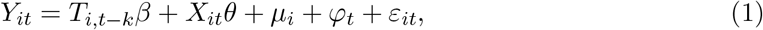

where *Y*_*it*_ denotes one of the two alternative outcome variables as measured in country *i* at time *t*; *T*_*i,t-k*_ is one of our two dichotomous treatment variables indicating whether a country participates in an IMF programme or whether it has implemented an IMF-mandated privatisation reform, lagged by *k* ∈ {1, 5, 10} years to allow for dynamic effects to manifest; *X*_*it*_ is a vector of control variables; *µ*_*i*_ captures time-invariant country-specific effects; *ϕ*_*t*_ measures time-fixed effects; and *ε*_*it*_ is a stochastic error term. Our principal quantity of interest is *β*, which is a causal effect parameter to be estimated. A common approach to estimating *β* is through the use of two-way fixed effects models that isolate variation within units over time whilst controlling for aggregate time trends. This has the virtue of eliminating all bias induced by time-invariant confounders. However, some countries might be more likely to seek the IMF’s assistance than other countries, and this might generate a spurious association between structural adjustment programmes and our outcomes of interest that cannot be addressed through the sole use of a fixed effects model.

The possibility of such a spurious association can be more intuitively illustrated as follows. Consider two doctors, A and B. Doctor A is a highly skilled clinician who, on the basis of a strong record, is only assigned patients whose treatment other doctors find too difficult or challenging and whose survival chances are low. Doctor B, on the other hand, is less competent and is therefore only assigned easier cases. If ‘success’ is defined in terms of patient survival, a naive comparison of the success rates of doctors A and B can lead to the faulty conclusion that doctor B is more competent than doctor A since the former’s patients have a higher marginal chance of survival. Such a conclusion fails to take into account the severity of the disease that each doctor is treating: since doctor A only treats the sickest patients, his or her success rate is bound to be low compared to doctor B, who only works with easily treated patients. In a similar vein, the IMF can be viewed as a doctor who intervenes across the world to treat (financially) ailing patients (countries), but caution is warranted in interpreting subsequent health outcomes without controlling for other factors that make some units more likely to receive the treatment than others (that is, pre-treatment ‘disease severity’). Previous research has indeed shown that failing to account for such bias in the study of IMF programmes almost invariably leads to meaningless results (21–23).

One way to address this concern is to adjust for as many confounders (*X*) as possible in our model. However, in an observational study of such a complex matter, it is hard to know whether all sources of confounding are adequately addressed. A different and more compelling solution is to construct an instrumental variable, *Z*, which is correlated with the treatment, *T*, but uncorrelated with any other variables in the causal system, thereby isolating quasi-random variation in *T*. For an intuitive understanding of this concept, consider the following example. Suppose we are interested in the causal effect of alcohol consumption on liver disease. We could simply compute the association between these two variables in a population but we are likely to be concerned that the corresponding effect estimate is biased, especially if there is a third unobserved factor that influences both alcohol consumption and liver disease. Such a factor might include an underlying propensity to health destructive behaviour or broader social determinants of health that drive such behaviours. To avoid this problem, we might exploit (e.g., regional) variation in the price of alcohol as an instrumental variable: the price of alcoholic beverages will affect drinking behaviour — but it is unlikely to affect liver disease other than through its effect on drinking behaviour. In other words, it is not correlated with other confounding factors and thus isolates what is known as ‘exogenous’ variation in the independent variable.

In our case, as per Figure 1, an instrumental variable (*Z*) is a variable that is predictive of the intervention of interest (*T* = IMF programmes) but that is uncorrelated with unmeasured confounders (*U*). These two criteria are known as the ‘relevance criterion’ (the instrument needs to be relevant to, i.e., have an impact on, the treatment) and the ‘exclusion criterion’ (the instrument must not be associated with other variables in the causal system under consideration), respectively. To obtain such an instrument, we follow recent methodological advances in the study of structural adjustment (21, 22) by adopting an instrument derived from the Fund’s annual budget constraint, as measured by the number of countries with an IMF programme in a given year (23). The IMF’s annual budget constraint should meet the two key criteria for an instrumental variable: it is clearly related to IMF programmes, but there is no reason to suspect that it is correlated with country-specific health profiles. In other words, whether or not the IMF is experiencing liquidity problems is not a function of the state of health in any given client country (just as the price of alcohol should not depend on any given individual’s propensity to drink). Moreover, previous research has shown that the IMF is more likely to impose harsher loan conditions when it faces liquidity concerns, regardless of the client in question (21–23). This allows us to specifically investigate ‘high-dose’ interventions by the Fund. In more technical terms, our identification strategy relies on a compound instrument derived from the interaction between the country-specific average exposure to IMF programmes over the sample period and the Fund’s annual budget constraint. This instrument has been carefully evaluated in the extant literature (21, 22). Further technical methodological details are provided in the Supplementary information.

**Figure 1:**
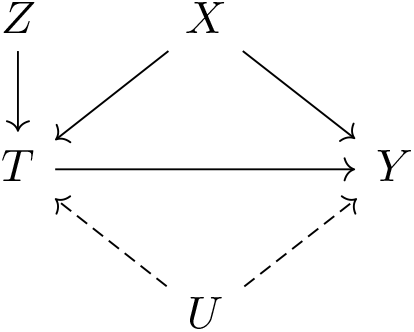
Causal graph depicting the effect of the treatment variable (*T*) on the outcome (*Y*), identified via a compound instrument (*Z*), net of both measured covariates (*X*) and unmeasured confounders (*U*).

Given that we cannot empirically verify whether or not the instrument is strictly exogenous, the persistence of unmeasured residual confounding is possible. To address this concern, we also conduct a simple yet comprehensive non-parametric sensitivity analysis that allows us to quantify the amount of unmeasured confounding that would in theory be required to eliminate our estimated treatment effect 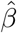. As per Figure 1, let *U* denote an unmeasured confounder. Then the bias factor, ℬ, is defined as the difference between 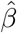 and what 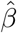 would have been had we controlled for *U* as well. Assuming *U* is binary, we define

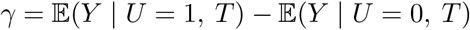

as the net effect of the unmeasured confounder on the outcome and

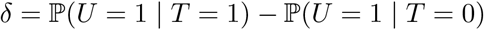

as the difference in the prevalence of the unmeasured confounder between the treatment and control groups. Then the bias factor is the product of these two sensitivity parameters: ℬ = *γ* · *δ*(24, 25). In assessing the sensitivity of our model coefficients to unmeasured confounding, we ask how large *γ* would have to be in order to reduce our estimated effect size 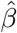 to zero. We address this question by visualising how ℬ changes as the two sensitivity parameters (co-)vary across a range of possible values.

## Results

Figure 2 displays our baseline results. As seen in the top half of the figure, we find that IMF programmes as a whole exert a substantively large impact both on mortality rates from respiratory infections and TB per 100,000 population, notably in the short (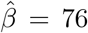 excess deaths, 95% CI: 42–110) and medium (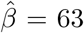 excess deaths, 95% CI: 20–106) term. In the long run, the effect dissipates somewhat and is harder to identify precisely, as evidenced by smaller effect sizes and widening confidence intervals (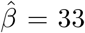 excess deaths, 95% CI: 4–62). As seen in the bottom half of the figure, this aggregate effect appears to be driven by IMF-mandated privatisation reforms, which generate large excess death burdens from communicable diseases (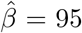 excess deaths after 1 year, 95% CI: 39–151). Also in this case, we find that the effect weakens after a decade (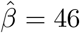 excess deaths after 10 years, 95% CI: −6–98).

**Figure 2:**
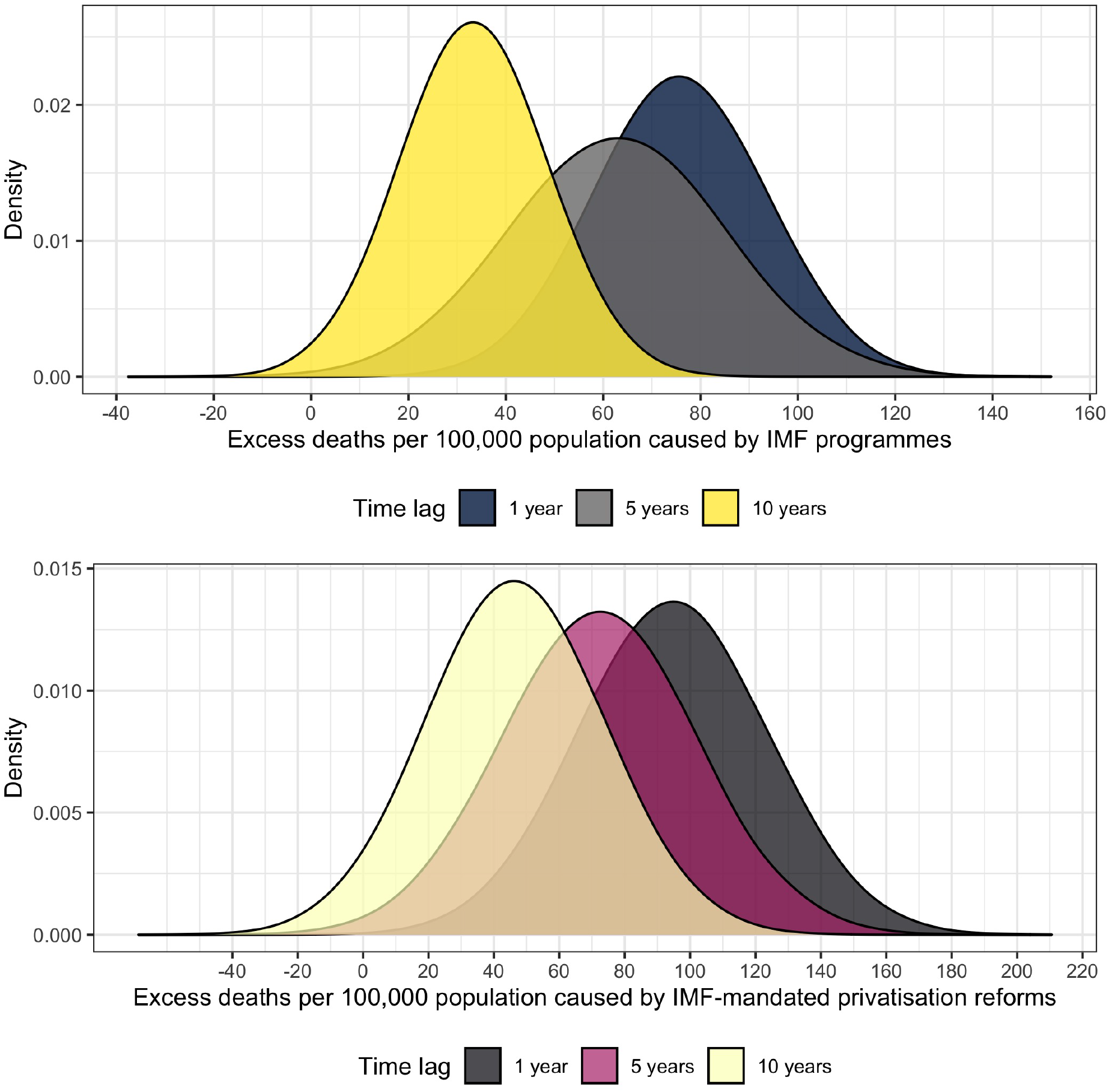
The figure visualises the estimated excess mortality burden from respiratory infections and tuberculosis per 100,000 population caused by IMF programmes as a whole (top) and IMF-mandated privatisation reforms (bottom). The estimates are derived from two-way fixed effects instrumental variable regression models in which within-country changes in mortality rates across units with and without IMF programmes or privatisation conditionalities are calculated. First differences in the outcome variable are then used to estimate excess death rates.

To assess any confounding from observed variables, we check the robustness of our parameter to additional control variables. To avoid multicollinearity and the loss of too many observations at once due to missing data, we add and remove these controls one by one and inspect the corresponding change in the treatment coefficient. To avoid unnecessary clutter, we limit our sensitivity analysis to the short-term effects identified above and omit all but key quantities of substantive interest (i.e., the estimated treatment effects). As displayed in Table 2, we find that structural adjustment remains a robust predictor of our outcome variable. The greatest attenuation in the estimated baseline treatment effects occurs when controlling for foreign reserves — which is an important predictor of selection into IMF programmes (23). As shown in the last row of the table, we also find evidence that health systems are an important mechanism through which these adjustments impact health, with estimates being attenuated by nearly one half when controlling for the number of hospital beds per 1,000 population. This is not the case for privatisation reforms, the coefficient for which remains stable despite increasing estimation uncertainty. We note, however, that the hospital beds per capita models are based on a substantially reduced sample size (total *N* = 2, 517) due to missing data and should therefore be interpreted with some caution.

**Table 2:** Instrumented two-way fixed effects control models

| CONTROL VARIABLE | IMF<sub>*t*-1</sub> COEFFICIENT | PRIVATISATION<sub>*t*-1</sub> COEFFICIENT |
| --- | --- | --- |
| Log of GDP per capita | 63<br>(24, 102) | 80<br>(8, 152) |
| Financial crisis | 77<br>(42, 112) | 95<br>(40, 150) |
| Foreign reserves | 47<br>(18, 76) | 86<br>(17, 155) |
| Democracy | 68<br>(35, 101) | 87<br>(30, 144) |
| Egalitarian democracy | 87<br>(30, 108) | 87<br>(18, 156) |
| Coup d'état | 75<br>(30, 120) | 76<br>(9, 143) |
| UNGA voting alignment | 73<br>(40, 106) | 89<br>(32, 146) |
| Female education | 79<br>(46, 112) | 97<br>(42, 152) |
| Hospital beds per 1,000 population | 31<br>(13, 49) | 93<br>(-1, 187) |

NOTES: The outcome variable is the age-standardised mortality rate from respiratory infections and tuberculosis per 100,000 population. Each row is a separate two-way fixed-effects regression wherein the effect of IMF programmes or IMF-mandated privatisation reforms on the outcome is adjusted for the control variable listed in the first column. All models are also adjusted for country- and time-fixed effects. The two treatment variables, lagged by one year, are instrumented as described in the METHODS section. 95% confidence intervals derived from standard errors consistent with heteroskedasticity, serial correlation, and unit clustering are shown in parentheses below each parameter estimate.

Given the observational nature of our study, the persistence of unmeasured residual confounding is possible. To address this concern, we conduct a simple non-parametric sensitivity analysis that allows us to quantify the amount of unmeasured confounding that would in theory be required to eliminate our estimated short-run causal effects, as described above. The results of this analysis are visualised in Figures 3 and 4. The Y-axis quantifies the net effect of *U* on the outcome variable that would be required to completely eliminate the estimated causal effect of structural adjustment programmes. In light of our instrumented treatment variable, we believe it is plausible that the amount of residual confounding remains small and that the difference in prevalence of any confounder between treatment and control groups should be minimal. As such, the most likely magnitude of *δ* would be at the lower end of the X-axis in Figure 3. If *δ* = 0.1, *U* would have to cause over 750 excess communicable disease deaths per 100,000 population to nullify the effect of IMF programmes. Even higher net effects, exceeding 900 excess deaths per 100,000, would be required to nullify the impact of privatisation conditionalities, as seen in Figure 4. Also at higher values of *δ*, an inordinate amount of unmeasured confounding would be needed to cast serious doubt on our effect estimates.

**Figure 3:**
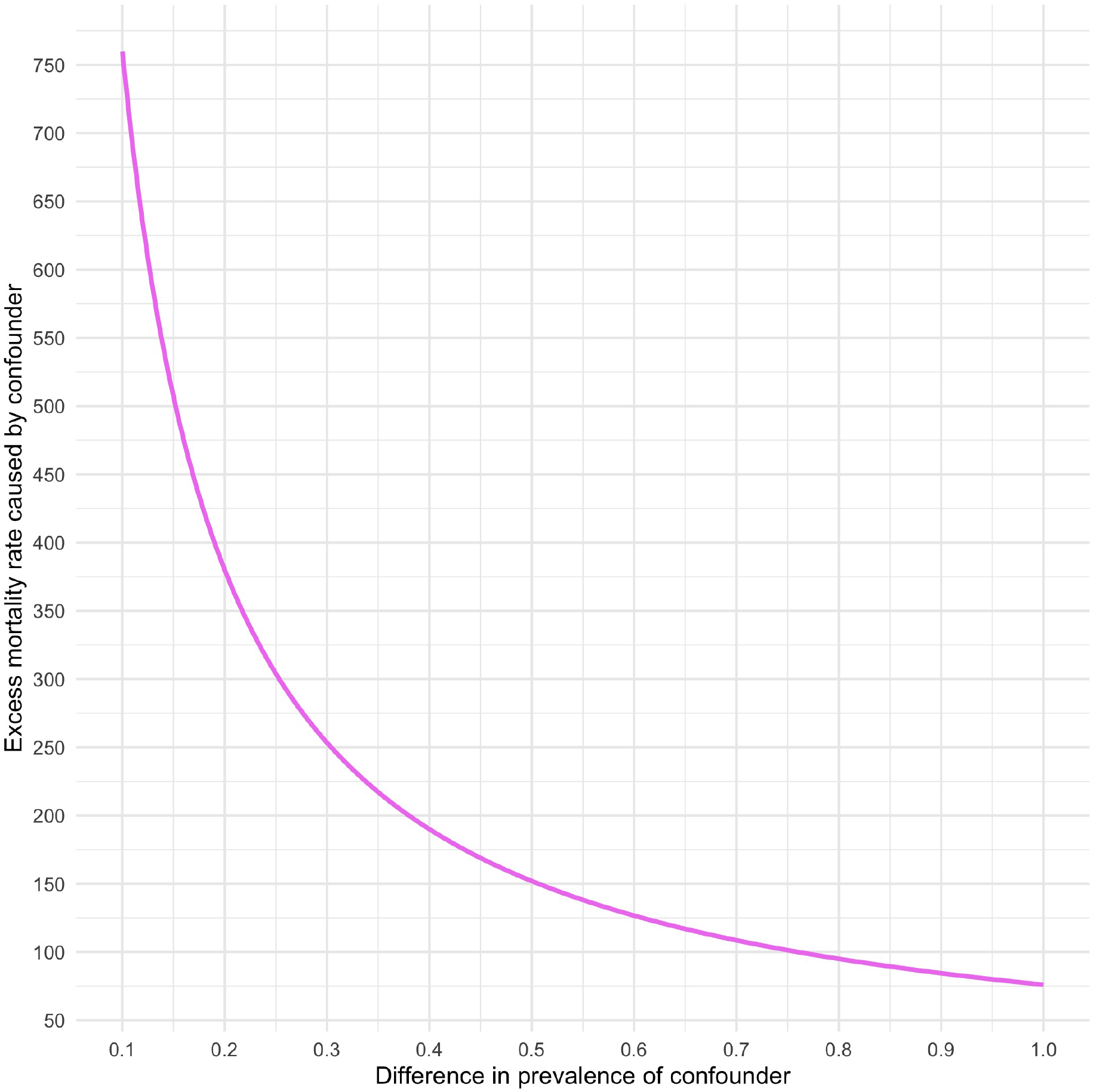
Sensitivity analysis plot to assess residual confounding of the estimated effect of IMF programmes on mortality rates from respiratory infections and tuberculosis as per the top half of Figure 2. Values on the solid lines would completely eliminate the estimated effect of IMF programmes. Values above the plotted curves would reverse the sign of the estimated effects.

**Figure 4:**
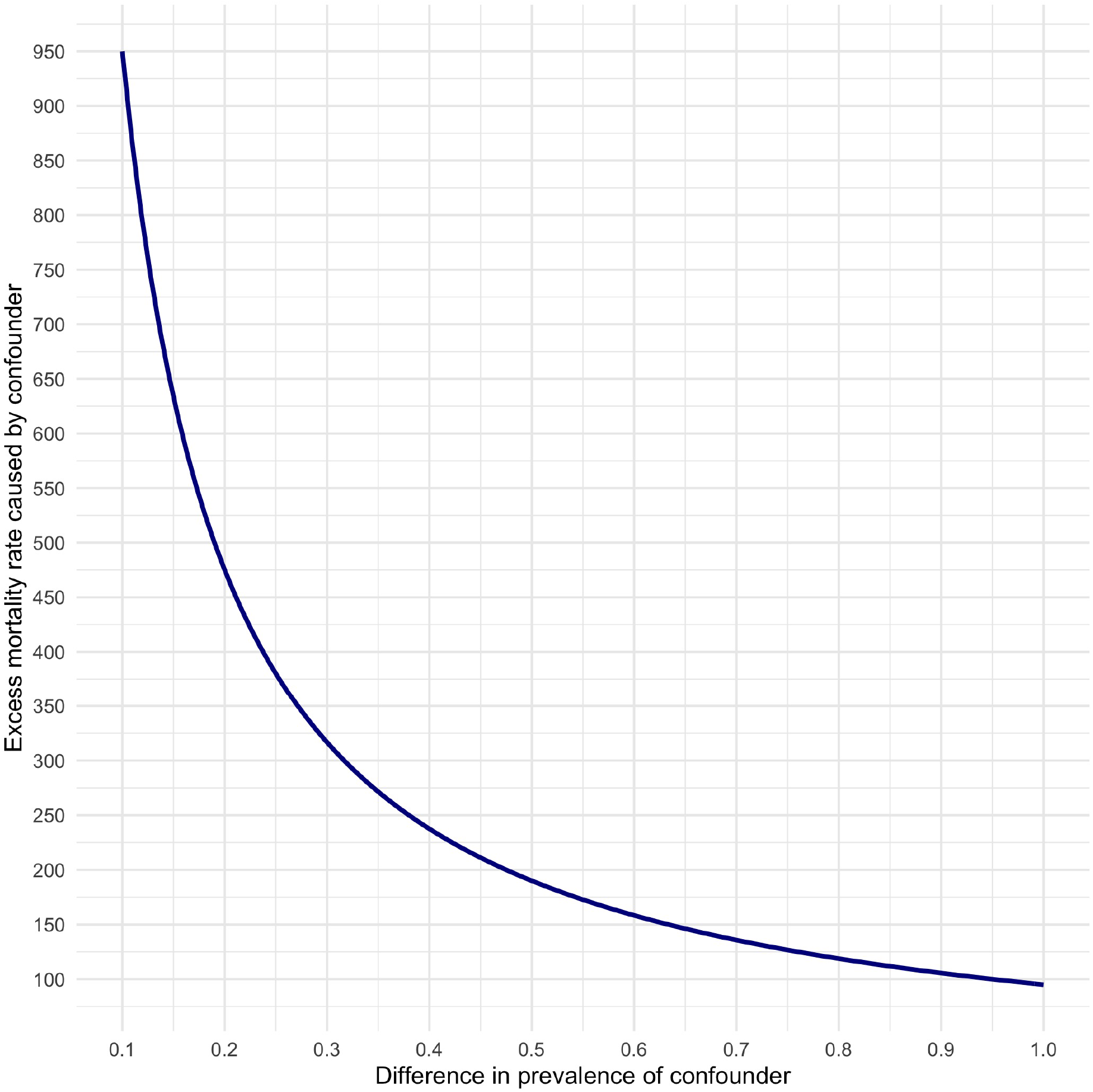
Sensitivity analysis plot to assess residual confounding of the estimated effect of IMF-mandated privatisation reforms on mortality rates from from respiratory infections and tuberculosis as per the bottom half of Figure 2. Values on the solid lines would completely eliminate the estimated effect of privatisation reforms. Values above the plotted curves would reverse the sign of the estimated effects.

## Concluding discussion

Our analysis provides novel evidence from previously unavailable cross-national panel data, linking the IMF’s policy interventions to poor health outcomes. We corroborate earlier studies and hypotheses surrounding this topic, yet we offer new empirical insights. Our main finding is that IMF programmes, and IMF-mandated privatisation reforms in particular, increase avoidable burdens of illness and death from respiratory infections and TB. Although we are unable to specify the specific mechanisms by which the estimated causal effects take place, we find that a substantial portion of the aggregate effect seems to be mediated by how IMF programmes impact local health systems, as measured by the number of available hospital beds per 1,000 population. This is in line with the extant literature, which suggests that IMF-mandated reductions in health and public sector spending affect the quality of primary care in addition to exerting durable influence on the social determinants of health (1). For lack of data, we are unable to probe the mechanisms any further, though we note that the rapid privatisation of state-owned enterprises has previously been linked to turbulent labour market conditions, high levels of social insecurity and stress, and weaker public institutions (9, 10). Such insights lend credence to our findings.

We note that the observational nature of our analysis precludes any guarantee of strictly unbiased model estimates. However, the sensitivity analysis suggests that an unusual amount of unmeasured confounding would be required to cast serious doubt on our substantive findings. We thus consider our results as providing an important empirical basis for future policy-making and research. We therefore conclude that structural adjustment programmes, as currently designed and implemented, are harmful to population health by increasing global communicable disease burdens and must be rethought and reformed.

## Data Availability

Available from lead author upon request.

## Supplementary information

Technically, our identification strategy relies on a compound instrument derived from the interaction between the country-specific average exposure to IMF programmes over the sample period and the Fund’s annual budget constraint, approximated by the number of countries with an IMF programme in a given year (23). This instrument meets the relevance criterion insofar as the IMF is likely to impose more stringent loan conditions when facing liquidity concerns. Previous research has indeed shown that the IMF’s budgetary constraint is predictive of harsher loan conditions, regardless of the client country (21, 23). The proposed instrument also meets the exclusion criterion insofar as the Fund’s aggregate annual budget constraints are independent of any given country, such that unit-specific shocks that deviate from a country’s long-run average exposure to structural adjustment result from a treatment assignment mechanism that is orthogonal to (i.e., uncorrelated with) any given country’s potential (or counterfactual) outcomes. In other words, the outcome of interest in countries with varying propensities to participate in IMF programmes will not be affected by changes in the Fund’s budgetary constraint other than through the impact of structural adjustment. Our identification strategy is visually summarised in Figure 1, where the conditional independence assumption obtains by deploying *Z* as a source of exogenous variation in *T* that helps us identify a local average treatment effect. Thus, the joint usage of an instrumental variable and of country- and time-fixed effects provides a rigorous framework for causal identification.

We thus obtain a two-stage regression model with the following selection equation:

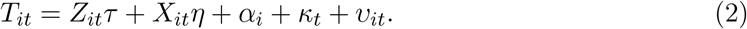

We then re-specify the model in equation (1) as follows, with 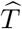 being a vector of fitted values from equation (2):

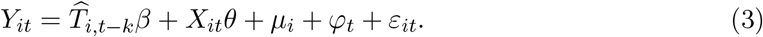

To empirically assess the strength of the chosen instrument, we compare the model in equation (2) to a restricted first-stage regression in which the effect *τ* of *Z* on *T* is set to be null, obtaining a *χ*^2^ test statistic of 53.641 on 1 degree of freedom (*p <* 0.001). Our alternative instrument targeting privatisation conditionalities also passes the required significance threshold, with a *χ*^2^ test statistic of 40.464 on 1 degree of freedom (*p <* 0.001). Hence, in both cases, *Z* comfortably satisfies convention criteria for strong instruments, meaning they do predict the treatment. We control for the endogenous relation between *T* and *Y* potentially induced by any time-invariant propensity of countries with a prior health disadvantage to select into IMF programmes by adjusting for country-fixed effects, whereas year-fixed effects help account for broader time trends that affect all countries simultaneously. All variance estimators are robust with respect to serial autocorrelation, heteroskedasticity, and country-level clustering effects. All analyses are conducted in R, version 4.0.2.

